# Milestone Development in Genetic Conditions from SFARI Registries

**DOI:** 10.1101/2021.04.07.21254183

**Authors:** Jordan Wickstrom, Cristan Farmer, LeeAnne Green Snyder, Andrew R. Mitz, Stephan J. Sanders, Somer Bishop, Audrey Thurm

## Abstract

**Background:** Several recent initiatives have sought to better understand characteristic behaviors of rare genetic conditions associated with autism spectrum disorder (ASD). The onset of developmentally expected skills, such as walking and talking, serve as readily quantifiable aspects of the behavioral phenotype. The goals of this study were to describe attainment of major motor and language milestones in genetic conditions implicated in the etiology of ASD and other neurodevelopmental disorders and to compare those phenotypes to idiopathic ASD.

**Methods:** Participants ages 3 years and older were drawn from two Simons Foundation Autism Research Initiative-funded registries with consistent phenotyping protocols. Inclusion criteria consisted of a confirmed genetic diagnosis for one of 16 specific genetic conditions (Simons Searchlight), and absence of pathogenic genetic findings in idiopathic ASD controls (SPARK). Parent-reported age of acquisition of three motor and two language milestones was described and quantified as on-time or delayed relative to normative expectations.

**Results:** Delay was more common among participants with rare genetic conditions than idiopathic ASD for all milestones. Compared to the idiopathic ASD group, the median odds of delay among the genetic groups were 8.3 times (IQR 5.8-16.3) higher for sitting, 12.4 times (IQR 5.3-19.5) higher for crawling, 26.8 times (IQR 7.7-41.1) higher for walking, 2.7 times (IQR 1.7-5.5) higher for single words, and 5.7 times (IQR 2.8-18.3) higher for combined words.

**Conclusion:** Delays in major developmental milestones, particularly in motor skills, may be among the earliest clues that developmental processes may be differentially affected in specific genetically defined conditions versus a behaviorally defined disorder such as idiopathic ASD.

---

Several recent initiatives have emerged to better describe behaviors characteristic of genetic conditions implicated in the etiology of autism spectrum disorder (ASD) and related neurodevelopmental disorders (NDDs). Large-scale collaborations (e.g., Simons Simplex Collection; Fischbach & Lord, 2010; Simons VIP Consortium, 2012) have made available larger cohorts of individuals with specific genetic diagnoses, allowing for a genetics-first approach to examining phenotypes. Defining the phenotypic landscape of these genetic conditions is important for improving identification and clinical management. Additionally, given their established association with ASD, characterizing these conditions may promote understanding of the etiological mechanisms underlying ASD-related phenotypes (Sanders et al., 2019).

Information regarding attainment of developmental milestones, such as walking and talking, is one way to quantify very early aspects of the phenotype. Extensive data on such developmental milestones are available for the general population (Sheldrick et al., 2019; Størvold, Aarethun, & Bratberg, 2013; Taanila, Murray, Jokelainen, Isohanni, & Rantakallio, 2005; World Health Organization, 2006). Developmental milestone attainment, particularly in motor and language skills, has also been a focal point in ASD research. Among children with ASD, early motor milestones are generally achieved within normal age limits (Bishop, Thurm, Farmer, & Lord, 2016; Havdahl et al., 2020; Matson, Mahan, Kozlowski, & Shoemaker, 2010), while the onset of language milestones is more variable (Eigsti, de Marchena, Schuh, & Kelley, 2011; Mayo, Chlebowski, Fein, & Eigsti, 2013; Mitchell et al., 2006). Behavioral phenotype data regarding early development (“developmental phenotypes”) from recently identified rare genetic conditions associated with ASD are beginning to emerge (Arnett et al., 2020; Bernier et al., 2017; Chawner et al., 2019; Hinton et al., 2013; Winders, Wolter-Warmerdam, & Hickey, 2019). Evidence from a phenotype-first perspective indicates that among children with ASD, those with identifiable rare genetic conditions walk later than those without known genetic conditions (Bishop et al., 2017).

The goal of the current study was to extend the limited literature on developmental phenotypes in multiple recently-identified genetic conditions implicated in ASD (1q21.1 deletion, 1q21.1 duplication, 16p11.2 deletion, 16p11.2 duplication, *ADNP, ASXL3, CSNK2A1, DYRK1A, GRIN2B, MED13L, PACS1, PPP2R5D, SCN2A, SLC6A1, STXBP1, SYNGAP1*), using contemporaneously and systematically collected data regarding motor and language milestone acquisition (Simons VIP Consortium, 2012). We compare these developmental phenotypes to those of a sample of children with ASD with no identified genetic condition (SPARK Consortium, 2018). Based on limited reports on milestone attainment in these genetic conditions in the literature, we expected to observe extensive delays relative to idiopathic ASD.

## Methods

### Cohorts

#### Simons Searchlight

Simons Searchlight, an initiative launched by the Simons Foundation Autism Research Initiative (SFARI) (previously known as Simons Variation in Individuals Project) (Simons VIP Consortium, 2012), is comprised of groups of people with rare genetic variants implicated in ASD. While nearly all participants have developmental disabilities of varying severity, not all participants have ASD. Individuals with a genetic diagnosis on the Simons Searchlight Gene List (https://www.simonssearchlight.org/research/what-we-study/) of 148 gene changes and 20 copy number variants (CNVs) enrolled in the registry from 2014 to September 2020. Participants were recruited worldwide by the Simons Searchlight community website (https://www.simonssearchlight.org) as well as with social media outreach, clinical referrals, and other means of web recruitment (e.g., http://www.clinicaltrials.gov) following diagnosis of one of the genetic conditions in Simons Searchlight. The protocol consists of developmental and behavioral surveys in an online study portal, a telephone interview to obtain medical history and administer the Vineland Adaptive Behavior Scales (Sparrow, Cicchetti, & Balla, 2005; Sparrow, Cicchetti, & Saulnier, 2016), and the collection of biospecimens. Files from Version 7 were downloaded on September 29, 2020 from SFARI.org, and included the 1q21.1, 16p11.2, and single gene mutation datasets.

#### SPARK

The SPARK registry is the largest existing genetic study of ASD (SPARK Consortium, 2018). Individuals from the U.S. with a reported professional ASD diagnosis and their family members are eligible for enrollment in SPARK, regardless of age or genetic status. Families are recruited by 31 sites across the U.S. using an extensive web recruitment strategy. Similar to Simons Searchlight, medical and developmental history, current behavioral data, and biospecimens were collected remotely through an online portal beginning in late 2015. Version 4 was downloaded on April 30, 2020 from SFARI.org.

### Sample Selection

The Supplemental Information contains a detailed description of how data were combined, as well as a flow diagram of sample selection (Figure S1). From both cohorts (i.e., Simons Searchlight and SPARK), participants for the current study were probands at least 3 years of age with verified genetic results, and at least one valid response for any of the five developmental milestones of interest. The minimum age was selected because normative acquisition of the milestones considered in these analyses occurs before the age of 3 years (Centers for Disease Control, June 10, 2020; Sheldrick et al., 2019; World Health Organization, 2006). This allowed for the interpretation of milestones which were reportedly “not yet achieved” as “delayed.”

Cohort-specific inclusion criteria were as follows. Simons Searchlight groups containing at least 15 cases with available walking milestone data were included in the genetic conditions sample, for a total of 479 participants across 16 conditions: four CNVs (1q21.1 deletion, 1q21.1 duplication, 16p11.2 deletion, 16p11.2 duplication) and twelve single genes (*ADNP, ASXL3, CSNK2A1, DYRK1A, GRIN2B, MED13L, PACS1, PPP2R5D, SCN2A, SLC6A1, STXBP1, SYNGAP1*). Simons Searchlight participants both with and without reported ASD diagnoses were included. Probands from the SPARK sample were included in the idiopathic ASD group (n=3,506) if they underwent genetic analyses and were found to have no pathogenic single gene or CNV events strongly associated with ASD based on SNP genotyping array, whole exome sequencing, and review by the SPARK medical genetics committee (Feliciano et al., 2019). Adults who independently enrolled in Simons Searchlight and SPARK were not included in this study, because reports of milestone attainment are only provided by caregivers of dependent children or adults.

### Measures

Upon enrolling in either registry, participants were invited to complete a standard set of questionnaires. All questionnaires for participants in the current study were completed by parents or caregivers. For a list of exact variables and datasets used for each registry, please refer to Figure S2. Whereas specific measures used for medical history and other data collection varied by registry (see subsections below), the developmental milestone and demographic data were collected via a Background History form in both registries.

The primary outcomes included ages of milestone attainment for sitting, crawling, walking, using single words, and using combined words. For each milestone, respondents chose one of the following: attainment age in months (selected from 1-84 months), achieved after age 7 years, or not yet achieved. However, the response options for the milestone questions changed slightly during the data collection period; the option of “not yet achieved” was introduced in 2018. Prior to this, parents of children who did not acquire a skill may have left the age of milestone attainment blank. To the extent that this is true, the observed data are biased toward children who had acquired a milestone.

For this reason, we explored options for partially imputing missing data on milestone acquisition using scores from the Vineland-II Adaptive Behavior Scales Survey Interview Form (Sparrow et al., 2005). The Vineland-II is a semi-structured interview that can be used across the lifespan. It measures adaptive functioning in four domains (with corresponding subdomains): Communication (Expressive, Receptive, Written), Daily Living (Personal, Domestic, Community), Socialization (Interpersonal Relationships, Play and Leisure Time, Coping Skills), and, in children under age 7, Motor Skills (Gross, Fine). Where available, scores from the Vineland-II were used to estimate whether a participant with missing milestone acquisition data had acquired walking and single words (see Supporting Information for a detailed explanation). Item-level data were not available, so estimated minimum raw scores compatible with the ability to walk (Gross Motor = 27) and use single words (Expressive Language = 19) were used to determine whether or not a participant had attained the milestone. Attainment scores were imputed for 4% of the sample for walking and 14% of the sample for single words. Raw scores less than the threshold were used to code the milestone as “not yet achieved.” If the raw scores exceeded the threshold, the milestone was coded as “achieved, age unknown.”

Finally, “delay” in the acquisition of milestones was operationalized using normative data. Specifically, motor milestones were considered to be delayed if they occurred after 8 months for sitting, 12 months for crawling, and 16 months for walking, based on the 97^th^ percentiles (8.4, 12, and 16, respectively) reported for normative data (World Health Organization, 2006). Language milestones were defined as delayed after 12 months for single words and 24 months for combined words based on the Act Early CDC Recommendations (Centers for Disease Control, June 10, 2020). Milestones which were not attained were considered delayed for the purpose of analysis. For descriptive purposes, the degree of delay was categorized as attainment occurring within 6 months or beyond 6 months (including those who never attained) of the expected age of acquisition.

#### Simons Searchlight

Parent-reported diagnoses of ASD, Intellectual Disability (ID), Seizure Disorder/Epilepsy, and gestational age were drawn from the medical history interview. Parents or caregivers also participated in the Vineland-II with licensed genetic counselors over the phone at the time of the medical history interview. Raw scores on the Expressive Communication and Gross Motor subdomains at first administration of the Vineland-II were used to guide the imputation of some missing milestone data, as described above.

#### SPARK

Parent-reported diagnosis of ID, Seizure Disorder/Epilepsy, gestational age, and additional demographics (i.e., ancestry) were drawn from registration, a background history form, and a medical screening survey.

### Statistical Analysis

The goals of this study were primarily descriptive. However, it was necessary to quantify the degree of difference in milestone acquisition between the genetic conditions and the idiopathic ASD sample. For the sake of clinical interpretability, we operationalized the outcome as on-time or delayed acquisition of a given milestone as described above. Given the uniform delay amongst some of the genetic conditions, we used the penalized likelihood-based method Firth logistic regression, using the logistf package for R version 4.0.2 (Heinze, Ploner, & Jiricka, 2020). Absolute (rate) and relative (odds ratios; OR) probabilities of delay, based on genetic condition, are provided alongside 95% confidence intervals. To further describe the patterns of onset, age of acquisition for those who achieved a milestone prior to age 7 years is summarized for each group using the median and interquartile range. This was possible only for those who acquired the milestone prior to 7 years due to the response options on the form. Proportions of the sample are also presented for those who acquired the skill after age 7 years or at an unknown age, did not acquire the skill, or had missing data regarding acquisition of that skill.

#### Missing Data

Non-response occurred for 179 participants (4% sample) on one milestone (Simons Searchlight n=46, SPARK n=133), 102 participants (3% sample) on two milestones (Simons Searchlight n=52, SPARK n=50), and 74 participants (2% sample) on three or four milestones (Simons Searchlight n=35, SPARK n=39). We also imposed one logical constraint on the data that created missing data: where the age of using combined words was younger than the age of using single words, we treated both as missing (Simons Searchlight n=5; SPARK n=15). To the extent possible, missing data were imputed as acquired/not acquired, as described above. Vineland-II data were available only for Simons Searchlight, and were not available for all participants who were missing data. Data were imputed for n=19 of 28 Simons Searchlight participants missing walking and n=69 of 80 missing single word. No other imputation of missing data was performed.

## Results

Sample demographics are provided in Table 1. The idiopathic ASD group had a higher proportion of males (82%) compared with the genetic conditions sample (53%). Half the samples maintained an annual household income of $81,000 or above (54% and 52%, respectively). For the genetic and idiopathic samples, the parent-reported prevalence of ASD was 37% and 100%, ID was 25% and 13%, Seizure Disorder/Epilepsy was 35% and 4%, and premature birth was 12% and 10%, respectively.

**Table 1.**
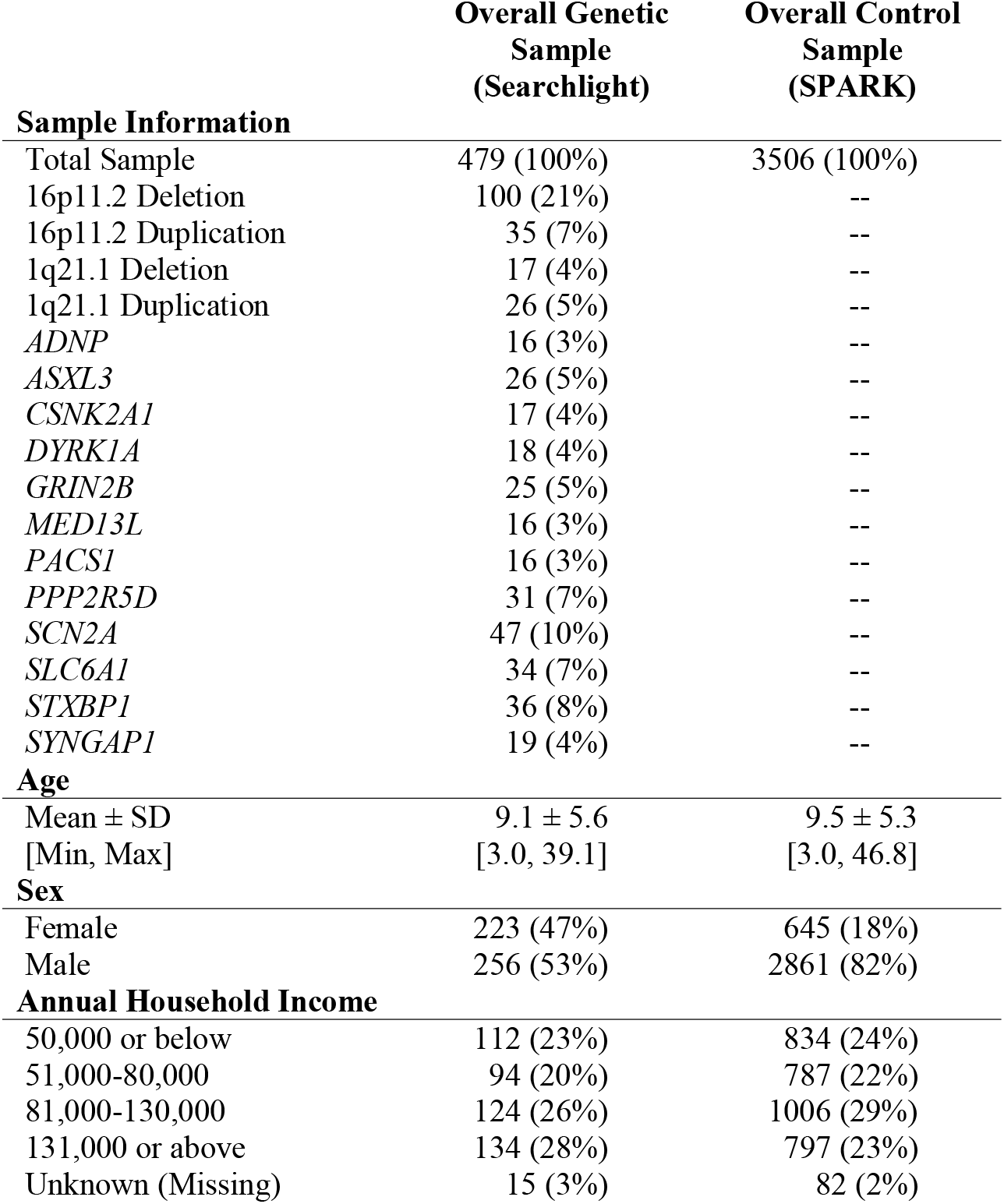
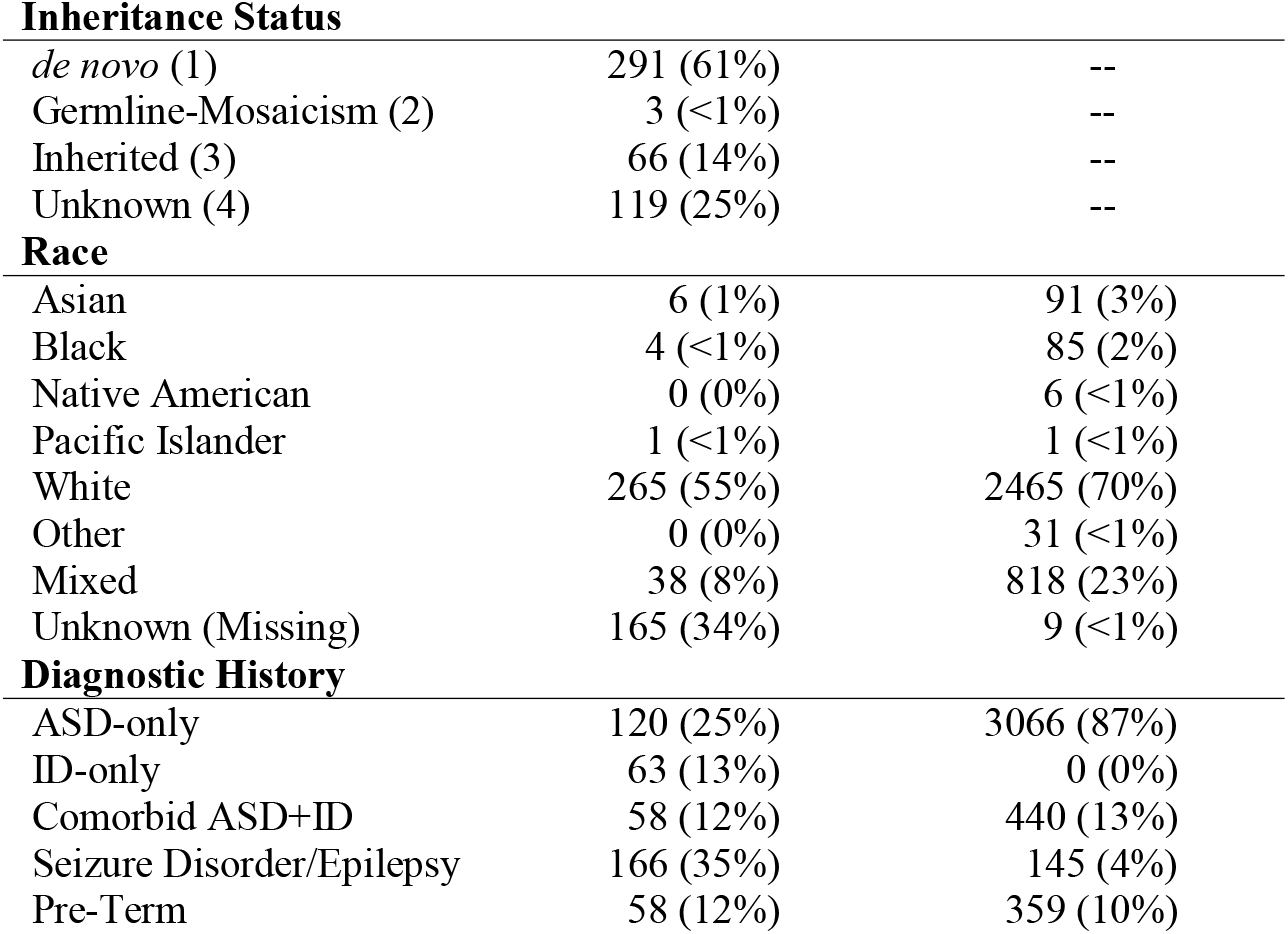
Sample demographics are provided for all genetic conditions in Simons Searchlight combined and the idiopathic ASD group from SPARK.

Descriptive statistics (quartiles and proportions) for milestone acquisition are provided in Table 2. Means and standard deviations for each milestone, if attained, are provided in Table S1. Overall, probands with genetic conditions were more likely to exhibit both lack of attainment and delayed attainment of milestones than the idiopathic ASD comparison group. Delays were most pronounced for motor milestones. Compared to the idiopathic ASD group, the median odds of delay for the genetic condition groups were higher by 8.3 times (IQR 5.8-16.3) for sitting, 12.4 times (IQR 5.3-19.5) for crawling, and 26.8 times (IQR 7.7-41.1) for walking. For the language milestones, the median odds of delay for the genetic condition groups were higher than the idiopathic ASD group by 2.7 times (IQR 1.7-5.5) for single words and 5.7 times (IQR 2.8-18.3) for combined words. The rate and extent of delay in milestone attainment is illustrated in Figure 1.

**Table 2.**
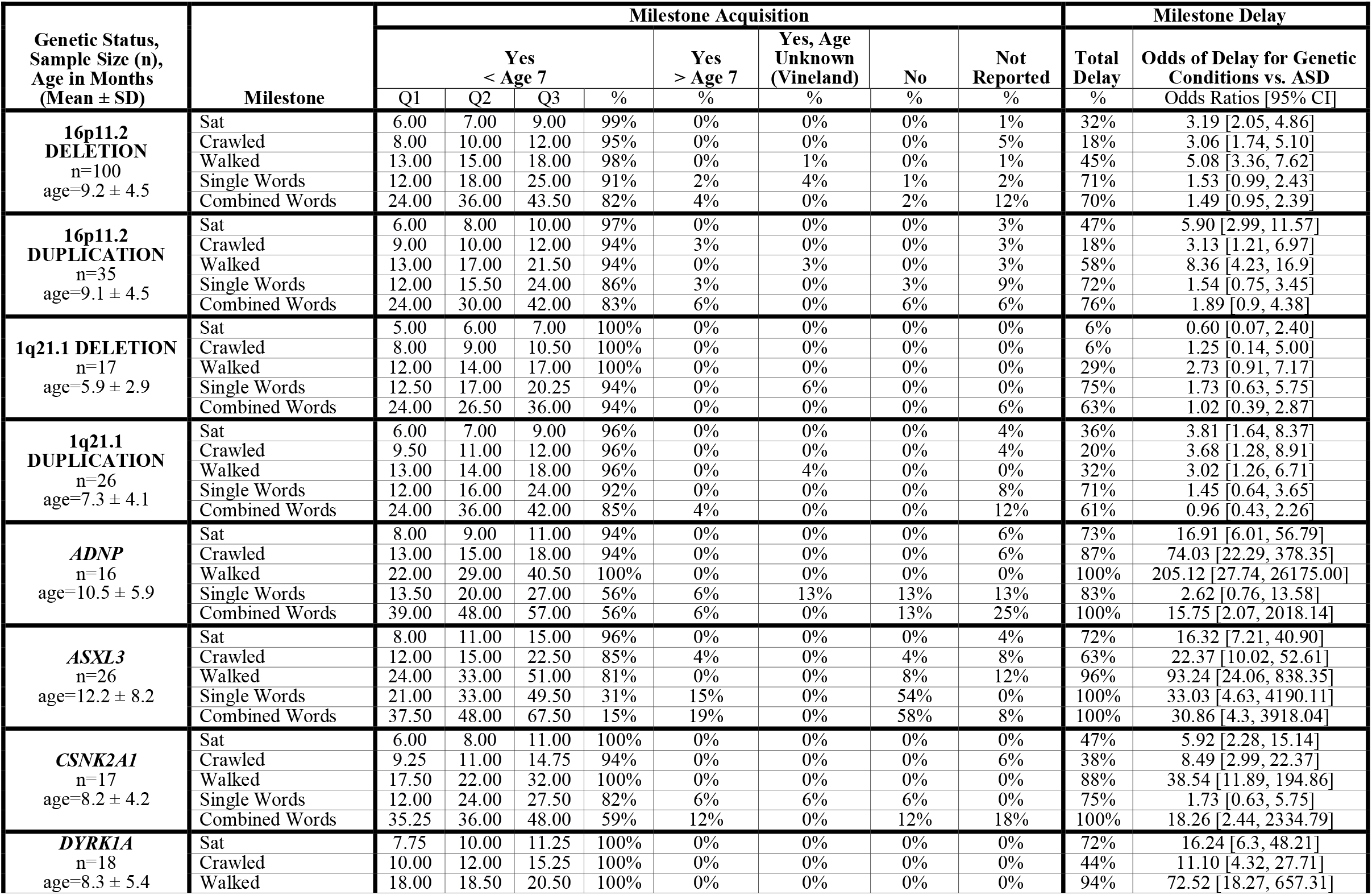
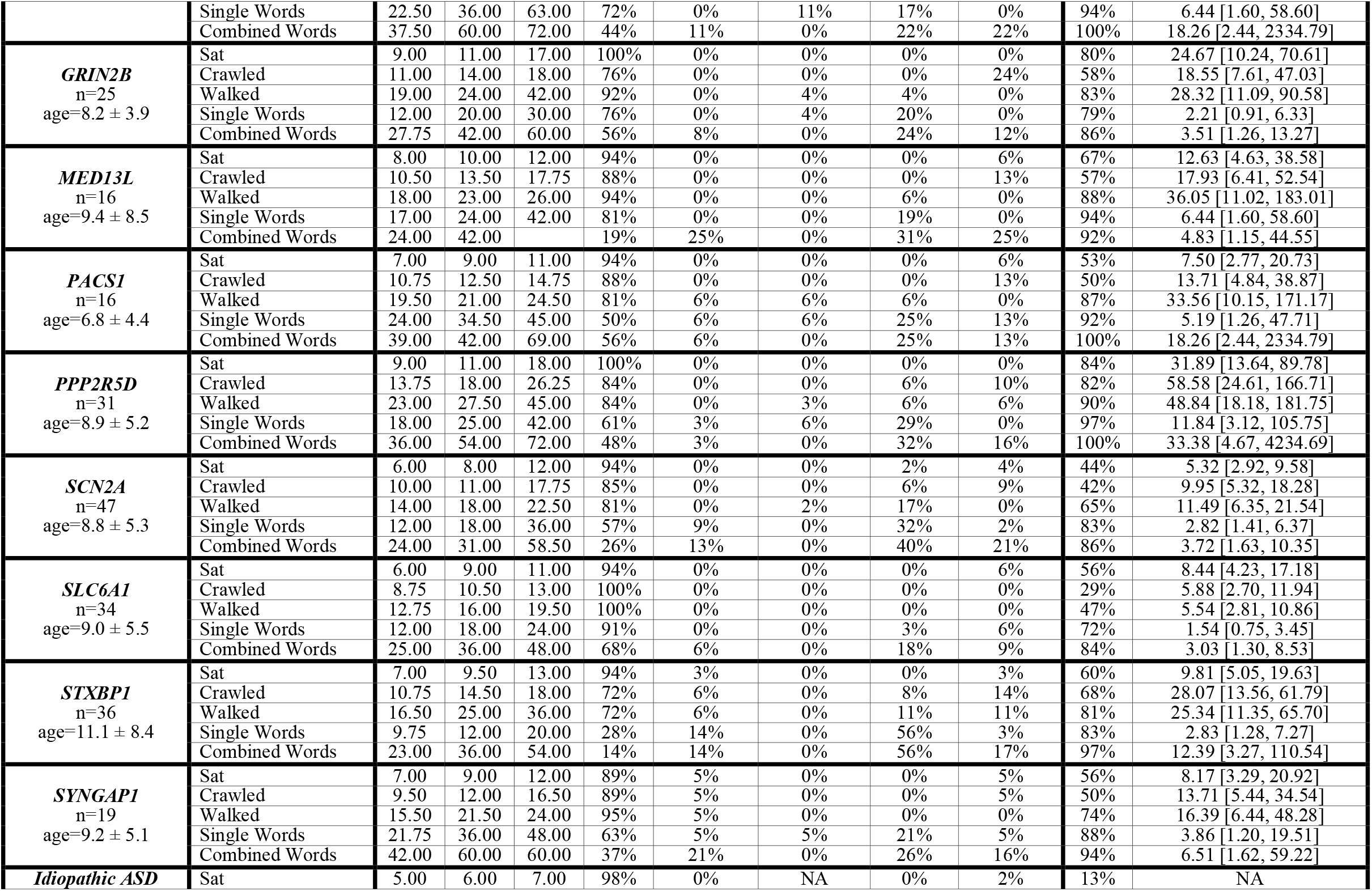
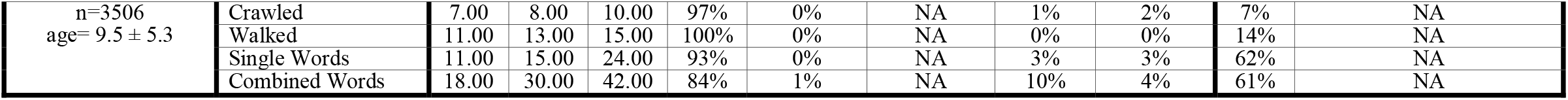
Quartiles for milestone acquisition, as well as proportion of the sample, are provided for those who achieved a milestone prior to age 7. Proportions of the sample are also presented for those who acquired the skill after age 7 or at an unknown age (age is unknown because Vineland scores were used to replace missing data where possible for walking and single words), did not acquire the skill, or had no reported data. Proportions of the sample that exhibited delays for each milestone are provided, as well as the odds ratios and confidence intervals for the odds of delay, for each respective genetic condition in comparison to the idiopathic ASD (SPARK) group.

**Figure 1.**
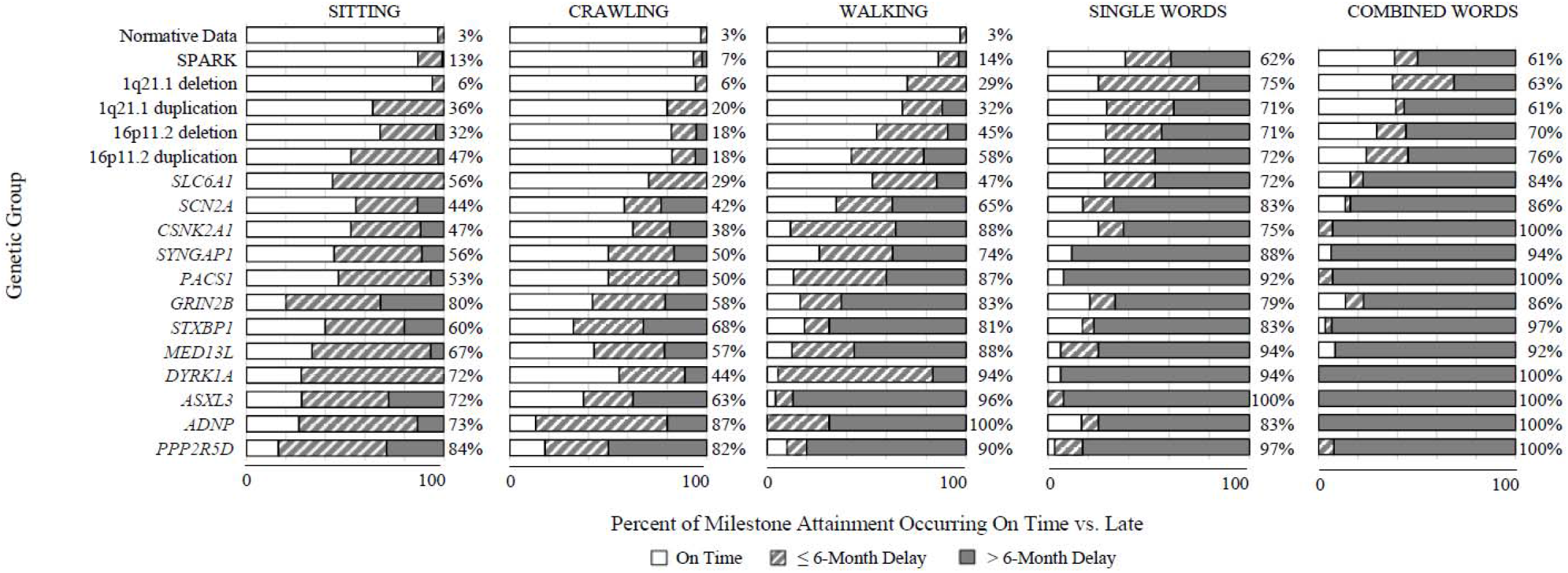
For each genetic group, the percent of milestone attainment is provided for sitting, crawling, walking, and single and combined words. This figure indicates the percentage of on-time versus late attainment, with late attainment divided by delays of up to six months versus beyond six months (including those who never attained the skill). Delays were defined as >8 months for sitting (WHO), >12 months for crawling (WHO), >16 months for walking (WHO), >12 months for single words (CDC), and >24 months for combined words (CDC). Total percentages for late attainment are provided in the right margin of each bar graph.

As shown in Table 2 and Figure 1, at least 50% of the sample was delayed in sitting for 10 genetic conditions, crawling for eight conditions, and walking for 12 conditions. All groups, including idiopathic ASD, had a rate of delay ≥50% for language milestones. Eight genetic conditions (*ADNP, ASXL3, GRIN2B, MED13L, PACS1, PPP2R5D, STXBP1*, and *SYNGAP1*) had delays for *all* milestones in at least half the sample.

## Discussion

This study examined developmental phenotypes across multiple rare genetic conditions, several of which were recently recognized, and compared them to a group identified through genetic analyses as idiopathic ASD. This study extends previous work in the Simons Simplex Collection (Bishop et al., 2017; Buja et al., 2018) by examining multiple early developmental milestones (sitting, crawling, walking, single-word talking, combined-word talking) from systematically-collected data on selected rare genetic conditions associated with ASD, and considering the genetic conditions individually rather than in aggregate. By comparing developmental phenotypes across specific genetic conditions to that of an idiopathic ASD group, we were able to add to a growing literature about diverse patterns of very early motor and language skill acquisition among probands with genetic conditions that are associated with higher rates of ASD and other NDDs. As anticipated, the early developmental milestone profiles of probands with genetic conditions were marked by more extensive delays (including lack of attainment) than the idiopathic ASD group, especially for the motor milestones.

While phenotypic data are just now becoming available from some of the genetic conditions reported here (Arnett et al., 2020; Berg, Palac, Wilkening, Zelko, & Schust Meyer, 2020; Hanly, Shah, Au, & Murias, 2020), the availability of systematically collected developmental and other behavioral phenotypic data from both the Simons Searchlight and SPARK registries allowed for cross-registry comparisons. Our focus on developmental milestones augments the available literature on cross-sectional cognitive and psychiatric profiles by documenting the earliest manifestations of phenotypes which may cascade into lifelong neurodevelopmental conditions. Examination of key motor and language milestones indicates that most children with the 16 genetic conditions reported here exhibit delays in both of these domains. In particular, varying patterns of delays in motor milestones were found across groups, with greater delays on average found in the single gene conditions compared to the CNVs included in this study. Of note, compared to single gene conditions, microarray testing for CNVs has been available longer, costs less, is more widely available, and is more established as a primary genetic test, all of which may have contributed over time to ascertainment of milder phenotypes in the population. Language delays on the other hand, tended to be consistently delayed across all genetic conditions, although there was much more variability in the complete lack of attainment of language milestones in the single gene conditions (3-56% single words; 12-58% combined words) compared to the CNVs or idiopathic ASD group (0-3% for single words, 0-10% for combined words). The genetic groups with the largest proportion of language delays also had the largest proportion of motor delays, which is consistent with previous literature suggesting that these systems are strongly related (Ghassabian et al., 2016; Libertus & Hauf, 2017).

The difference between idiopathic ASD and the genetic conditions was most obvious for the motor milestones. While this study is not the first to note early motor impairments associated with rare genetic mutations (Buja et al., 2018), or generally intact motor milestones of children diagnosed with idiopathic ASD (Bishop et al., 2016; Havdahl et al., 2020; Matson et al., 2010; Ozonoff et al., 2008), it extends the findings of others by documenting this pattern across multiple genetic condition groups. These results reveal the great extent and variability of delays, and support the idea that severe motor delays may be indicative of genetic etiologies (Bishop et al., 2017; Satterstrom et al., 2020). Our description of delays using established norms and demarcation of delay exceeding 6 months (including those who never attained) illustrates the magnitude of delay in the selected genetic conditions, and suggests that exacerbated motor delays are a likely predictor of these types of genetic abnormalities. By contrast, delays in language milestones were ubiquitous across all the genetic conditions, as well as the idiopathic ASD group, confirming previous research in ASD that delays in early language are not specific to probands with an identifiable genetic condition.

Single gene conditions, such as those included in this study, may be induced in animal and cell culture model systems, providing a mechanism to investigate pathogenesis of ASD-related phenotypes. Questions regarding whether these models can actually provide insight into *all* cases of idiopathic ASD, which models might be best-suited to this objective, and whether ASD neurobiology can be distinguished from broader developmental disabilities, have yet to be addressed. Answering these questions will require a more detailed understanding of the relationships between developmental delays and ASD; however, thus far both gene discovery and genotype-phenotype analyses support a model in which single gene mutations seem to vary in their contributions to developmental delay versus ASD (Satterstrom et al., 2020). Our results are consistent with this model. Single gene disorders with high rates of significant delays in early milestones (e.g. *PPP2R5D, ASXL3*) may be observed more frequently in developmental delay/ID cohorts than in ASD cohorts, while those with relatively lower (but still clinically significant) rates of delayed milestones (e.g. *SLC6A1, SCN2A)* may be observed at similar rates in developmental delay/ID cohorts *as well as* in ASD cohorts (Satterstrom et al., 2020). Of note, none of the genes observed at a higher rate in ASD than developmental delay (e.g., *PTEN, NRXN1*) from Satterstrom et al. (2020) were included in this study. With larger sample sizes and consistent ascertainment, developmental phenotype results across gene groups are likely to provide critical benchmarks to orient the results of model system analyses (Sanders et al., 2019).

## Limitations

A main limitation of this study is that the sample was drawn from registries based on self-referral, and small sample sizes precluded the inclusion of some groups (only 16 of the 29 genetic conditions with data currently available in Simons Searchlight are represented here). Further, participants in the Simons Searchlight registry were enrolled based on genetic testing, which could have been precipitated by delays in early milestones, as developmental delay is one of the main indications for genetic testing. These results may therefore reflect an ascertainment bias in the genetic condition group. These registries are not population-based, so it is possible that individuals with more severe phenotypes are not represented. In this way, the extent to which ascertainment strategies may affect these results likely varies by the different conditions and diagnoses. At present, we do not consider a multitude of factors that may be affecting variability within genetic subgroups, including inheritance and variant effect (e.g., missense vs. protein-truncating variant). In addition, the idiopathic ASD group included here may go on to receive a genetic diagnosis as new genetic discoveries are made. Still, a major strength of this study is that the idiopathic group was defined not by the lack of any “known genetic syndrome,” as is common in ASD research, but rather confirmed to not have significant genetic findings based on the current state of the field.

Given the high socioeconomic status and the distribution and missing data on race of the participants in the registries, these results may not be generalizable to the larger population of probands with these conditions. In addition, the age of milestone acquisition was not collected past the age of 7 years, and missing data were common, so the summary statistics for acquisition may be biased.

## Conclusions

For children with genetic conditions associated with ASD, findings here provide evidence that delays in motor and language milestones are among the early indicators of a diverging developmental trajectory. Motor delays may distinguish children with specific identifiable genetic etiologies from those with idiopathic ASD and should prompt early referral for genetic testing. Further, given its different profile, currently defined “idiopathic” ASD may reflect differing etiologies and mechanisms from rare genetic variants. Broader and deeper data are needed to investigate the possibility of distinguishing among genetic conditions on the basis of phenotypic profiles.

## Data Availability

Approved researchers can obtain the Simons Searchlight population dataset described in this study https://www.sfari.org/resource/simons-searchlight/ by applying at https://base.sfari.org. Approved researchers can obtain the SPARK population dataset described in this study https://www.sfari.org/resource/spark/ by applying at https://base.sfari.org.

## Abbreviations

(ASD): Autism Spectrum Disorder
(CNVs): Copy Number Variants
(ID): Intellectual Disability
(NDDs): Neurodevelopmental Disorders
(SPARK): Simons Foundation Powering Autism Research

## Acknowledgements

This research was supported (in part) by the Intramural Research Program of the NIMH (1ZICMH002961), an NIMH grant (U01 MH111662, SJS), and by an Autism Science Foundation (SJS, SB) grant. We are grateful to all of the families at the participating Simons Searchlight sites as well as the Simons Searchlight Consortium. We appreciate the opportunity to obtain access to Simons Searchlight phenotypic data on the SFARI Base. Approved researchers can obtain the Simons Searchlight population dataset described in this study https://www.sfari.org/resource/simons-searchlight/ by applying at https://base.sfari.org. In addition, we are thankful for the chance to gain access to the SPARK phenotypic data on the SFARI Base. We are grateful to all of the families in SPARK, the SPARK clinical sites and SPARK staff. Approved researchers can obtain the SPARK population dataset described in this study https://www.sfari.org/resource/spark/ by applying at https://base.sfari.org.

- What’s Known
  - There is an increasing number of genes and genetic conditions being identified that impart risk for autism spectrum disorder and other neurodevelopmental disorders.
  - Charting early developmental milestones in genetic conditions, based on norms acquired from typically developing children, provides valuable information regarding developmental phenotypes that characterize these conditions.
- What’s New
  - Compared to children with idiopathic autism spectrum disorder and typically developing children, children with the 16 rare genetic conditions reported here generally display more pronounced delays in early motor and expressive language milestones.
- What’s Relevant
  - Description of early developmental phenotypes in genetic conditions associated with autism spectrum disorder may be helpful for referral and identification, as well as for the search for mechanisms for potential interventions.

