## Supplemental Materials for "Milestone Development in Genetic Conditions from SFARI Registries"

**Vineland Scores as Proxy for Missing Data**

As a proxy for determining whether missing data could be treated as "not yet achieved" or "achieved age unknown," Vineland scores were used to augment missing data. Specifically, for age of attainment for walking, n=28 Simons Searchlight participants had missing data. Of those 28, n=19 had Vineland scores, and n=12 had gross motor raw scores ≤26, a score cut-off that can be interpreted as children being unable to walk at that time. Thus, for those 12 Simons Searchlight participants with gross motor raw scores ≤26, we treated as "not yet achieved," and for the remaining 7 participants >26 we treated as "achieved, age unknown." On the other hand, for age of attainment for single words, n=80 Simons Searchlight participants had missing data. Of those 80, n=69 had Vineland scores and n=54 had expressive language raw scores ≤18, a score cut-off that can be interpreted as children being unable to speak single words at that time. Thus, for those 54 Simons Searchlight participants with expressive language raw scores ≤18, we treated as "not yet achieved" for both language milestones (assuming that if a child is unable to speak single words, he/she is unable to speak combined words), and for the remaining 15 participants >18 we treated as "achieved, age unknown" for the single words milestone. Vineland scores were all within 11 months of the background history form completion, with the exception of three scores (17, 18 and 20 months).

**How Data Were Combined in Simons Searchlight and SPARK**

Simons Searchlight

1. For all 3 Searchlight Datasets (Simons_Searchlight_1q21.1_Dataset_v7.0, Simons_Searchlight_16p11.2_Dataset_v7.0, Simons_Searchlight_Single_Gene_Dataset_v7.0), from each of the level 1 subfolders (1q21.1 deletion, 1q21.1 duplication, 16p11.2 deletion, 16p11.2 duplication, ADNP, ARID1B, ASXL3, CHAMP1, CHD2, CHD8, CSNK2A1, CTNNB1, DYRK1A, FOXP1, GRIN1, GRIN2B, HIVEP2, HNRNPH2, MED13L, PACS1, PPP2R1A, PPP2R5D, SCN2A, SETBP1, SETD5, SLC6A1, STXBP1, SYNGAP1, TRIP12), all bghx_child.csv files were merged into a single .csv file. This .csv file also contained two additional groups (distal 16p11.2 deletion, distal 16p11.2 duplication) that were located in the level 1 subfolders for 16p11.2 deletion and duplication, respectively. This totaled n=897 participants across 31 groups. *Note: there were not any bghx_child.csv files for ANKRD11, MBD5, PTCHD1 or TBR1.*
2. In the single .csv file, applied participant inclusion/exclusion criteria:
    a. In the "age_at_eval" column:
        i. Kept participants 36 months and older
        ii. Eliminated participants under age 36 months or blank cells n=225
    b. Across the five columns "q2_sat_without_support_age", "q3_crawled_age", "q4_walked_age", and "q8_used_words_age", and "q11_combined_words_age":
        i. Kept participants who had data reported for at least 1/5 columns/ milestones
        ii. Eliminated participants who had no data for ANY of the 5 columns/ milestones: n=57
    c. In the "relationship_to_iip" column:
        i. Kept "Initially identified proband" participants
        ii. Eliminated "Siblings" and "Mothers" n=54
    d. In the "genetic_status_source" column:
        i. Kept "confirmed" participants

        ii. Eliminated “reported” or blank cells n=16
3. In the single .csv file, applied group inclusion/exclusion criteria:
    a. In the column “q4_walked_age” column:
        i. Kept groups who had responses for at least 15 participants
        ii. Eliminated groups who had data for less than 15 participants (n=111): ARID1B n=4, CHAMP1 n=10, CHD2 n=5, CHD8 n=7, CTNNB1 n=11, 16p11.2 distal deletion n=13, 16p11.2 distal duplication n=5, FOXPI n=6, GRIN1 n=5, HIVEP2 n=10, HNRNPH2 n=13, PPP2R1A n=2, SETBP1 n=13, SETD5 n=4, TRIP12 n=3
4. In the single .csv file, performed data cleaning:
    a. In the “sfari_id” column:
        i. Kept participants with unique identifiers
        ii. Eliminated participants with duplicate identifiers (this occurred for four identifiers; n=8)
5. After steps 1-4, the remaining 479 participants across 16 groups were included in the analysis.

<u>SPARK</u>

1. For the SPARK Dataset (SPARK_Collection_Version4), we started with a list (retrieved from SFARI representative) of 4,960 participants confirmed to have no identified genetic results (i.e., idiopathic ASD), and we merged the background_history-child.csv file with this list of participants into a single .csv file.
2. In the single .csv file, applied participant inclusion/exclusion criteria:
    a. In the “age_at_eval_months” column:
        i. Kept participants 36 months and older
        ii. Eliminated participants under age 36 months or blank cells n=1,422
    b. Across the five columns “sat_wo_support_age_mos”, “crawled_age_mos”, “walked_age_mos”, “used_words_age_mos”, and “combined_words_age_mos”:
        i. Kept participants who had data reported for at least 1/5 columns/ milestones
        ii. Eliminated participants who had no data for ANY of the 5 columns/ milestones: n=1,306
3. In the single .csv file, applied group inclusion/exclusion criteria:
    a. In the column “walked_age_mos” column:
        i. Kept group if had responses for at least 15 participants
        ii. Eliminated group if had data for less than 15 participants: n=0
4. In the single .csv file, performed data cleaning:
    a. In the “subject_sp_id” column:
        i. Kept participants with unique identifiers
        ii. Eliminated participants with duplicate identifiers: n=0
5. After steps 1-4, the remaining 3,506 participants were included in the analysis.

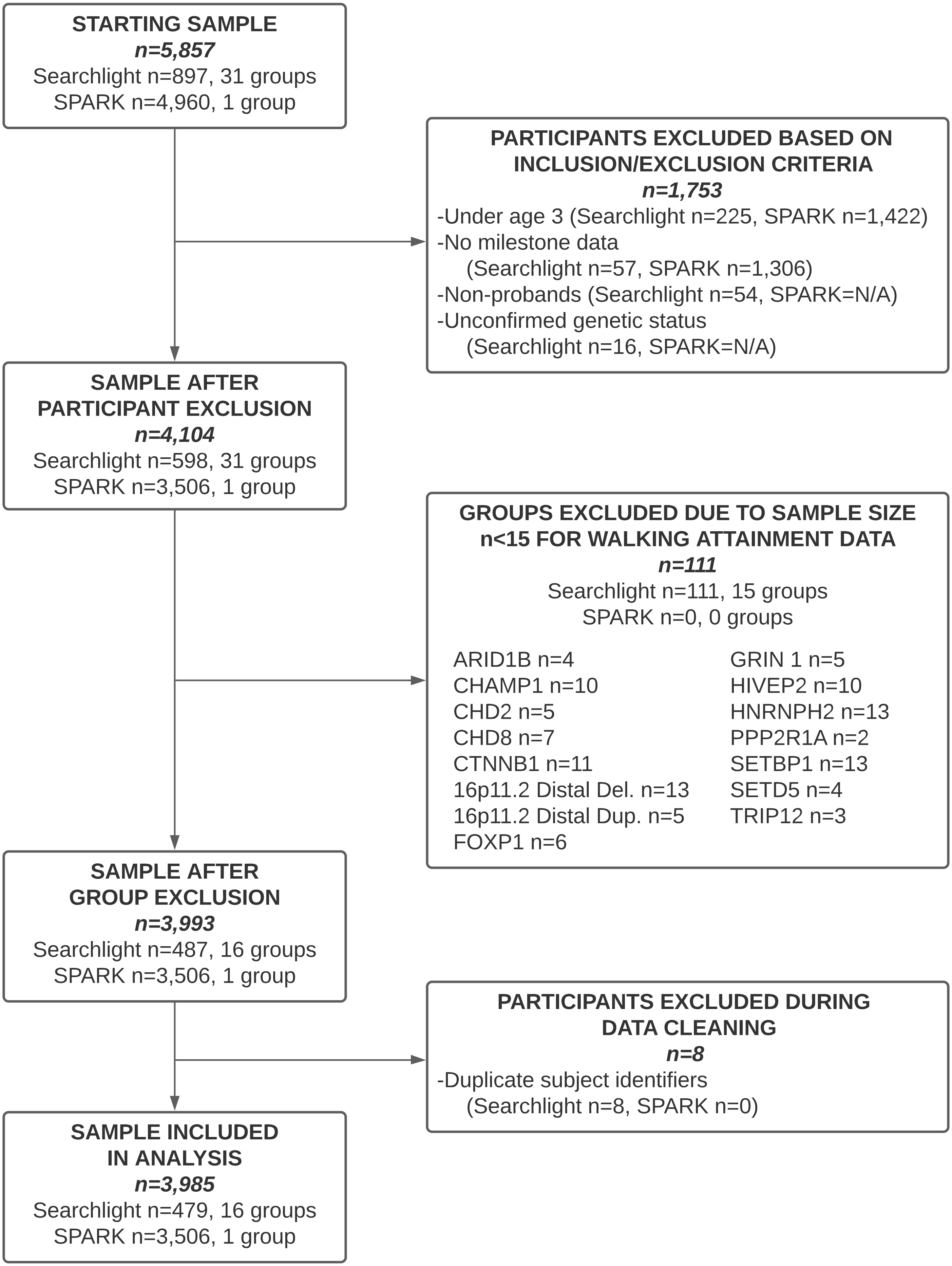


Figure S1. Flow chart of sample selection.

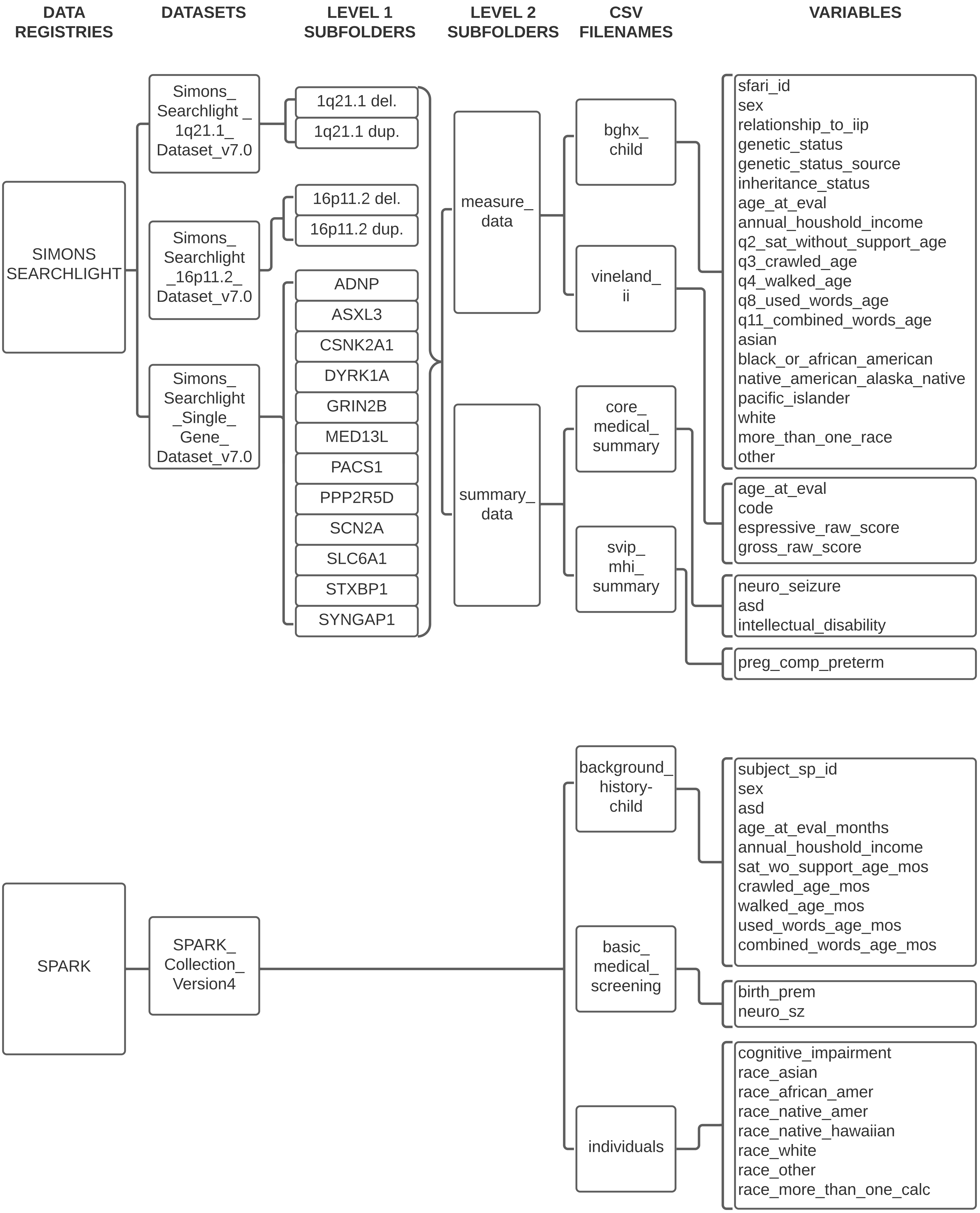


Figure S2. List of exact variables and their respective file locations used in the analysis from each registry.

| Genetic Status, Sample Size (n) and Age in Months (Mean ± SD) | Milestone | Attained < Age 7 | | | | | | | Attained > Age 7 | Attained Age Unknown | Not Yet Attained | Missing Data |
|---|---|---|---|---|---|---|---|---|---|---|---|---|
| | | N | Mean | Std. | Q1 | Q2 | Q3 | % | % | % | % | % |
| **16P11.2 DELETION** n=100 age=9.2 ± 4.5 | Sat | 99 | 8.06 | 3.15 | 6.00 | 7.00 | 9.00 | 99% | 0% | 0% | 0% | 1% |
| | Crawled | 95 | 10.57 | 4.11 | 8.00 | 10.00 | 12.00 | 95% | 0% | 0% | 0% | 5% |
| | Walked | 98 | 16.53 | 6.11 | 13.00 | 15.00 | 18.00 | 98% | 0% | 1% | 0% | 1% |
| | Single Words | 91 | 20.92 | 11.97 | 12.00 | 18.00 | 25.00 | 91% | 2% | 4% | 1% | 2% |
| | Combined Words | 82 | 35.17 | 15.08 | 24.00 | 36.00 | 43.50 | 82% | 4% | 0% | 2% | 12% |
| **16P11.2 DUPLICATION** n=35 age=9.1 ± 4.5 | Sat | 34 | 8.53 | 2.57 | 6.00 | 8.00 | 10.00 | 97% | 0% | 0% | 0% | 3% |
| | Crawled | 33 | 10.61 | 3.22 | 9.00 | 10.00 | 12.00 | 94% | 3% | 0% | 0% | 3% |
| | Walked | 33 | 16.94 | 4.78 | 13.00 | 17.00 | 21.50 | 94% | 0% | 3% | 0% | 3% |
| | Single Words | 30 | 19.13 | 10.14 | 12.00 | 15.50 | 24.00 | 86% | 3% | 0% | 3% | 9% |
| | Combined Words | 29 | 33.72 | 10.00 | 24.00 | 30.00 | 42.00 | 83% | 6% | 0% | 6% | 6% |
| **1Q21.1 DELETION** n=17 age=5.9 ± 2.9 | Sat | 17 | 6.29 | 2.31 | 5.00 | 6.00 | 7.00 | 100% | 0% | 0% | 0% | 0% |
| | Crawled | 17 | 9.06 | 2.56 | 8.00 | 9.00 | 10.50 | 100% | 0% | 0% | 0% | 0% |
| | Walked | 17 | 14.53 | 3.50 | 12.00 | 14.00 | 17.00 | 100% | 0% | 0% | 0% | 0% |
| | Single Words | 16 | 16.50 | 6.34 | 12.50 | 17.00 | 20.25 | 94% | 0% | 6% | 0% | 0% |
| | Combined Words | 16 | 27.94 | 7.78 | 24.00 | 26.50 | 36.00 | 94% | 0% | 0% | 0% | 6% |
| **1Q21.1 DUPLICATION** n=26 age=7.3 ± 4.1 | Sat | 25 | 7.92 | 2.47 | 6.00 | 7.00 | 9.00 | 96% | 0% | 0% | 0% | 4% |
| | Crawled | 25 | 11.44 | 2.74 | 9.50 | 11.00 | 12.00 | 96% | 0% | 0% | 0% | 4% |
| | Walked | 25 | 15.68 | 5.34 | 13.00 | 14.00 | 18.00 | 96% | 0% | 4% | 0% | 0% |
| | Single Words | 24 | 18.58 | 10.91 | 12.00 | 16.00 | 24.00 | 92% | 0% | 0% | 0% | 8% |
| | Combined Words | 22 | 35.23 | 14.36 | 24.00 | 36.00 | 42.00 | 85% | 4% | 0% | 0% | 12% |
| **ADNP** n=16 age=10.5 ± 5.9 | Sat | 15 | 10.47 | 3.76 | 8.00 | 9.00 | 11.00 | 94% | 0% | 0% | 0% | 6% |
| | Crawled | 15 | 16.73 | 5.48 | 13.00 | 15.00 | 18.00 | 94% | 0% | 0% | 0% | 6% |
| | Walked | 16 | 30.44 | 9.91 | 22.00 | 29.00 | 40.50 | 100% | 0% | 0% | 0% | 0% |
| | Single Words | 9 | 20.67 | 8.76 | 13.50 | 20.00 | 27.00 | 56% | 6% | 13% | 13% | 13% |
| | Combined Words | 9 | 47.33 | 9.22 | 39.00 | 48.00 | 57.00 | 56% | 6% | 0% | 13% | 25% |
| **ASXL3** n=26 age=12.2 ± 8.2 | Sat | 25 | 13.56 | 11.03 | 8.00 | 11.00 | 15.00 | 96% | 0% | 0% | 0% | 4% |
| | Crawled | 22 | 18.09 | 11.25 | 12.00 | 15.00 | 22.50 | 85% | 4% | 0% | 4% | 8% |
| | Walked | 21 | 37.24 | 18.92 | 24.00 | 33.00 | 51.00 | 81% | 0% | 0% | 8% | 12% |
| | Single Words | 8 | 36.38 | 18.73 | 21.00 | 33.00 | 49.50 | 31% | 15% | 0% | 54% | 0% |
| | Combined Words | 4 | 51.00 | 15.87 | 37.50 | 48.00 | 67.50 | 15% | 19% | 0% | 58% | 8% |
| **CSNK2A1** n=17 age=8.2 ± 4.2 | Sat | 17 | 9.06 | 4.04 | 6.00 | 8.00 | 11.00 | 100% | 0% | 0% | 0% | 0% |
| | Crawled | 16 | 14.00 | 8.12 | 9.25 | 11.00 | 14.75 | 94% | 0% | 0% | 0% | 6% |
| | Walked | 17 | 24.18 | 9.46 | 17.50 | 22.00 | 32.00 | 100% | 0% | 0% | 0% | 0% |
| | Single Words | 14 | 21.14 | 8.62 | 12.00 | 24.00 | 27.50 | 82% | 6% | 6% | 6% | 0% |
| | Combined Words | 10 | 41.10 | 9.39 | 35.25 | 36.00 | 48.00 | 59% | 12% | 0% | 12% | 18% |

| Genetic Status, Sample Size (n) and Age in Months (Mean ± SD) | Milestone | Attained < Age 7 | | | | | | | Attained > Age 7 | Attained Age Unknown | Not Yet Attained | Missing Data |
|---|---|---|---|---|---|---|---|---|---|---|---|---|
| | | N | Mean | Std. | Q1 | Q2 | Q3 | % | % | % | % | % |
| | Sat | 18 | 9.22 | 2.84 | 7.75 | 10.00 | 11.25 | 100% | 0% | 0% | 0% | 0% |
| **DYRK1A** | Crawled | 18 | 12.61 | 4.46 | 10.00 | 12.00 | 15.25 | 100% | 0% | 0% | 0% | 0% |
| n=18 | Walked | 18 | 19.33 | 5.85 | 18.00 | 18.50 | 20.50 | 100% | 0% | 0% | 0% | 0% |
| age=8.3 ± 5.4 | Single Words | 13 | 41.85 | 23.49 | 22.50 | 36.00 | 63.00 | 72% | 0% | 11% | 17% | 0% |
| | Combined Words | 8 | 57.75 | 18.12 | 37.50 | 60.00 | 72.00 | 44% | 11% | 0% | 22% | 22% |
| | Sat | 25 | 13.56 | 9.72 | 9.00 | 11.00 | 17.00 | 100% | 0% | 0% | 0% | 0% |
| **GRIN2B** | Crawled | 19 | 14.32 | 5.71 | 11.00 | 14.00 | 18.00 | 76% | 0% | 0% | 0% | 24% |
| n=25 | Walked | 23 | 28.83 | 16.80 | 19.00 | 24.00 | 42.00 | 92% | 0% | 4% | 4% | 0% |
| age=8.2 ± 3.9 | Single Words | 19 | 24.68 | 14.81 | 12.00 | 20.00 | 30.00 | 76% | 0% | 4% | 20% | 0% |
| | Combined Words | 14 | 42.79 | 17.80 | 27.75 | 42.00 | 60.00 | 56% | 8% | 0% | 24% | 12% |
| | Sat | 15 | 10.00 | 3.64 | 8.00 | 10.00 | 12.00 | 94% | 0% | 0% | 0% | 6% |
| **MED13L** | Crawled | 14 | 14.43 | 5.29 | 10.50 | 13.50 | 17.75 | 88% | 0% | 0% | 0% | 13% |
| n=16 | Walked | 15 | 21.87 | 7.59 | 18.00 | 23.00 | 26.00 | 94% | 0% | 0% | 6% | 0% |
| age=9.4 ± 8.5 | Single Words | 13 | 30.23 | 16.56 | 17.00 | 24.00 | 42.00 | 81% | 0% | 0% | 19% | 0% |
| | Combined Words | 3 | 46.00 | 24.25 | 24.00 | 42.00 | | 19% | 25% | 0% | 31% | 25% |
| | Sat | 15 | 8.53 | 3.74 | 7.00 | 9.00 | 11.00 | 94% | 0% | 0% | 0% | 6% |
| **PACS1** | Crawled | 14 | 12.50 | 5.83 | 10.75 | 12.50 | 14.75 | 88% | 0% | 0% | 0% | 13% |
| n=16 | Walked | 13 | 20.23 | 8.32 | 19.50 | 21.00 | 24.50 | 81% | 6% | 6% | 6% | 0% |
| age=6.8 ± 4.4 | Single Words | 8 | 32.38 | 12.85 | 24.00 | 34.50 | 45.00 | 50% | 6% | 6% | 25% | 13% |
| | Combined Words | 9 | 50.78 | 19.06 | 39.00 | 42.00 | 69.00 | 56% | 6% | 0% | 25% | 13% |
| | Sat | 31 | 14.55 | 10.12 | 9.00 | 11.00 | 18.00 | 100% | 0% | 0% | 0% | 0% |
| **PPP2R5D** | Crawled | 26 | 21.12 | 12.53 | 13.75 | 18.00 | 26.25 | 84% | 0% | 0% | 6% | 10% |
| n=31 | Walked | 26 | 32.92 | 18.16 | 23.00 | 27.50 | 45.00 | 84% | 0% | 3% | 6% | 6% |
| age=8.9 ± 5.2 | Single Words | 19 | 29.32 | 12.67 | 18.00 | 25.00 | 42.00 | 61% | 3% | 6% | 29% | 0% |
| | Combined Words | 15 | 54.27 | 17.25 | 36.00 | 54.00 | 72.00 | 48% | 3% | 0% | 32% | 16% |
| | Sat | 44 | 9.55 | 4.71 | 6.00 | 8.00 | 12.00 | 94% | 0% | 0% | 2% | 4% |
| **SCN2A** | Crawled | 40 | 14.05 | 6.67 | 10.00 | 11.00 | 17.75 | 85% | 0% | 0% | 6% | 9% |
| n=47 | Walked | 38 | 19.11 | 6.60 | 14.00 | 18.00 | 22.50 | 81% | 0% | 2% | 17% | 0% |
| age=8.8 ± 5.3 | Single Words | 27 | 24.04 | 16.54 | 12.00 | 18.00 | 36.00 | 57% | 9% | 0% | 32% | 2% |
| | Combined Words | 12 | 39.00 | 19.86 | 24.00 | 31.00 | 58.50 | 26% | 13% | 0% | 40% | 21% |
| | Sat | 32 | 8.94 | 2.79 | 6.00 | 9.00 | 11.00 | 94% | 0% | 0% | 0% | 6% |
| **SLC6A1** | Crawled | 34 | 10.82 | 3.00 | 8.75 | 10.50 | 13.00 | 100% | 0% | 0% | 0% | 0% |
| n=34 | Walked | 34 | 17.29 | 6.47 | 12.75 | 16.00 | 19.50 | 100% | 0% | 0% | 0% | 0% |
| age=9.0 ± 5.5 | Single Words | 31 | 20.94 | 10.74 | 12.00 | 18.00 | 24.00 | 91% | 0% | 0% | 3% | 6% |
| | Combined Words | 23 | 37.83 | 13.67 | 25.00 | 36.00 | 48.00 | 68% | 6% | 0% | 18% | 9% |

| Genetic Status, Sample Size (n) and Age in Months (Mean ± SD) | Milestone | Attained < Age 7 | | | | | | | Attained > Age 7 | Attained Age Unknown | Not Yet Attained | Missing Data |
|---|---|---|---|---|---|---|---|---|---|---|---|---|
| | | N | Mean | Std. | Q1 | Q2 | Q3 | % | % | % | % | % |
| **STXBP1** n=36 age=11.1 ± 8.4 | Sat | 34 | 12.85 | 12.78 | 7.00 | 9.50 | 13.00 | 94% | 3% | 0% | 0% | 3% |
| | Crawled | 26 | 15.04 | 6.78 | 10.75 | 14.50 | 18.00 | 72% | 6% | 0% | 8% | 14% |
| | Walked | 26 | 26.62 | 14.36 | 16.50 | 25.00 | 36.00 | 72% | 6% | 0% | 11% | 11% |
| | Single Words | 10 | 15.40 | 9.08 | 9.75 | 12.00 | 20.00 | 28% | 14% | 0% | 56% | 3% |
| | Combined Words | 5 | 38.00 | 16.49 | 23.00 | 36.00 | 54.00 | 14% | 14% | 0% | 56% | 17% |
| **SYNGAP1** n=19 age=9.2 ± 5.1 | Sat | 17 | 9.94 | 5.06 | 7.00 | 9.00 | 12.00 | 89% | 5% | 0% | 0% | 5% |
| | Crawled | 17 | 13.29 | 5.25 | 9.50 | 12.00 | 16.50 | 89% | 5% | 0% | 0% | 5% |
| | Walked | 18 | 20.78 | 6.51 | 15.50 | 21.50 | 24.00 | 95% | 5% | 0% | 0% | 0% |
| | Single Words | 12 | 36.50 | 20.82 | 21.75 | 36.00 | 48.00 | 63% | 5% | 5% | 21% | 5% |
| | Combined Words | 7 | 54.00 | 18.65 | 42.00 | 60.00 | 60.00 | 37% | 21% | 0% | 26% | 16% |
| **SPARK** n=3506 age=9.5 ± 5.3 | Sat | 3446 | 6.33 | 2.33 | 5.00 | 6.00 | 7.00 | 98% | 0% | NA | 0% | 2% |
| | Crawled | 3388 | 8.75 | 3.96 | 7.00 | 8.00 | 10.00 | 97% | 0% | NA | 1% | 2% |
| | Walked | 3494 | 13.56 | 4.61 | 11.00 | 13.00 | 15.00 | 100% | 0% | NA | 0% | 0% |
| | Single Words | 3276 | 19.64 | 12.63 | 11.00 | 15.00 | 24.00 | 93% | 0% | NA | 3% | 3% |
| | Combined Words | 2959 | 31.44 | 15.33 | 18.00 | 30.00 | 42.00 | 84% | 1% | NA | 10% | 4% |

Table S1. Sample sizes (N), means, and standard deviations are provided for all milestones achieved prior to age 7. Proportions of each group are indicated for each milestone according to those who achieved prior to age 7, after age 7, or at an unknown age (based on Vineland scores), for those who never attained the milestone, and for those unknown due to missing data.
